# The burden of patient healthcare activity in the first year of peritoneal dialysis

**DOI:** 10.1101/2024.02.19.24301896

**Authors:** Kirsty Crowe, Eleanor C Murray, Joan MacLeod, Jamie P Traynor, Peter C Thomson

**Author notes:** Correspondence: Dr Kirsty Crowe, Queen Elizabeth University Hospital, 1345 Govan Road, Glasgow, G51 4TF; 0141 451 6198; @croki_crowe.

## Abstract

**Background:** The healthcare-related burden associated with kidney replacement therapy (KRT) has not been characterised as comprehensively in peritoneal dialysis (PD) as in haemodialysis (HD) or kidney transplantation. This study aimed to capture the nature and extent of healthcare activity in the first year of PD therapy.

**Methods:** Retrospective analysis was undertaken on consecutive incident adult patients on PD between 1^st^ January 2015-31^st^ December 2019 in the Glasgow Renal and Transplant Unit. Healthcare-related activity was captured up to the first 365 days post-commencement of PD. Data was collected on renal service contact and activity relating to dialysis access, radiological investigation, and relevant infection episodes. Carbon mapping of healthcare activity was estimated using postcode data and previously published carbon footprint estimations.

**Results:** PD was initiated in 122 patients over the study period. Sixty-three patients (52%) transitioned to another KRT within 365 days of commencing PD. Patients had a mean 36.4 days (SD 22.7) of face-to-face contact days with renal services. This included a mean of 1.5 (SD 1.6) hospital admissions, with a median 5 (IQR 10.8) in-patient days. The estimated carbon footprint was 581kg CO2e/patient over the incident year. This included a median 207kg CO2e/patient for inpatient days and 26kg CO2e/patient for treatment of infections.

**Conclusions:** There is a significant burden of kidney-associated healthcare on patients commencing their first year of PD despite it being a home-based therapy. Estimates of carbon footprint indicate hotspots include patient travel and hospital admissions, and episodes of peritonitis; a full life cycle analysis is merited.

**Key Learning Points:** *What was known:*

- The characterisation of healthcare-related activity to be expected with peritoneal dialysis therapy is not as comprehensive as that for haemodialysis and kidney transplantation.
- Patient-centred realistic medicine requires knowledge of the patient journey and the cumulative impact of healthcare activity interactions.
- Healthcare activity burden mapping is required to determine the necessary carbon emission reductions for reducing the contribution to climate change.

*This study adds:*

- An illustration of the associated health-care activity burden on patients commencing peritoneal dialysis.
- An indication of the carbon footprint associated with peritoneal dialysis health-care activity in the first year of therapy.

*Potential impact:*

- Informative for service providers of peritoneal dialysis in anticipating care requirements and planning carbon reduction strategies.
- Data can help inform patient decision making when receiving education of kidney replacement therapies.

## Introduction

Peritoneal dialysis (PD) is the incident kidney replacement therapy (KRT) for 21% of adults with end-stage kidney disease in the UK[1]. PD is associated with better quality of life and a favourable cost-utility ratio compared to haemodialysis therapy, although has remained relatively static in prevalence, at around 5.5% of the total UK KRT population, since 2017[1][2]. Conveying the extent of the healthcare-related activity to be expected with KRT is essential for informed patient choice, but has not been characterised as comprehensively in PD as in haemodialysis (HD) or kidney transplantation[3][4]. Knowledge of the patient journey and the cumulative impact of renal service interactions is key to practising patient-centred realistic medicine, and essential when considering service configuration, resource deployment, and the targeting of carbon emission reductions necessary for healthcare services to reduce the contribution to climate change. This study aimed to capture the nature and extent of kidney-related healthcare activity in the first year of PD therapy.

## Materials and Methods

Consecutive incident adult patients on PD between 1^st^ January 2015-31^st^ December 2019 in the Glasgow Renal & Transplant unit were identified via a structured query language search of the Strathclyde Electronic Renal Patient Record (SERPR). The Glasgow Renal & Transplant unit is a tertiary renal centre providing PD services for a patient population of 1.6 million across two Scottish health boards (NHS Greater Glasgow & Clyde, and NHS Forth Valley). PD catheter insertion and initial PD training, delivered by PD specialist nurses, is undertaken at the tertiary centre, and follow-up clinic activity distributed between the centre and two peripheral district general hospital sites, depending on patient location. An ambulatory assessment area is available in the tertiary centre for rapid physician assessment and investigation of patients on an outpatient basis to avoid admission. PD catheter insertion is undertaken by a small team of surgeons affiliated with the renal unit. Technique varies between laparoscopic and open approach based on surgeon’s preference. There was no significant change in either staff members or insertion technique during the study period.

Demographic data including age, sex, postcode and primary renal diagnosis was extracted from SERPR. Retrospective analysis was undertaken on prospectively captured healthcare-related activity interrogated via SERPR and reports manually checked for accuracy. Healthcare-related activity was expressed per 1000 patient days and was captured up to the first 365 days post first PD catheter use for dialysis, censored by death or permanent switch in KRT modality unless the healthcare activity was directly related to PD (i.e. peritonitis related to PD catheter which remained in situ). The date for each healthcare activity was coded from 0 days to 365 post PD catheter use for dialysis. The patient cohort was sorted in order of number of days spent on PD in their first year, to facilitate a visual interpretation of temporal trends of modality switching and interaction with healthcare services.

Data were collected on all renal service interactions: hospital inpatient bed stays, renal ambulatory care visits, scheduled and unscheduled PD nurse and nephrologist telephone and in-person outpatient clinics and PD nurse home visit activity. Dialysis access activity, radiological investigation and data on relevant infection episodes – PD peritonitis, PD catheter infection, bloodstream infection, urinary tract infection - was also captured. Infection episodes were included if there were associated symptoms of infection and positive microbiological growth from tissue culture or in the case of PD peritonitis if a PD effluent white cell count >100/μL on microscopy (and > 50% polymorphonuclear leucocytes). All episodes of organism growth on microbiological blood cultures were included even if clinically treated as a contaminant result.

Carbon mapping of healthcare activity was estimated using postcode data and previously published carbon footprint estimations, as outlined in table 1. Pharmaceutical-related greenhouse gas emissions of empiric antibiotics for the infections listed above were included in the carbon calculations, but patients’ routine medications were not. Neither was the peritoneal dialysis itself included, only the additional healthcare activity related to KRT.

**Table 1.**
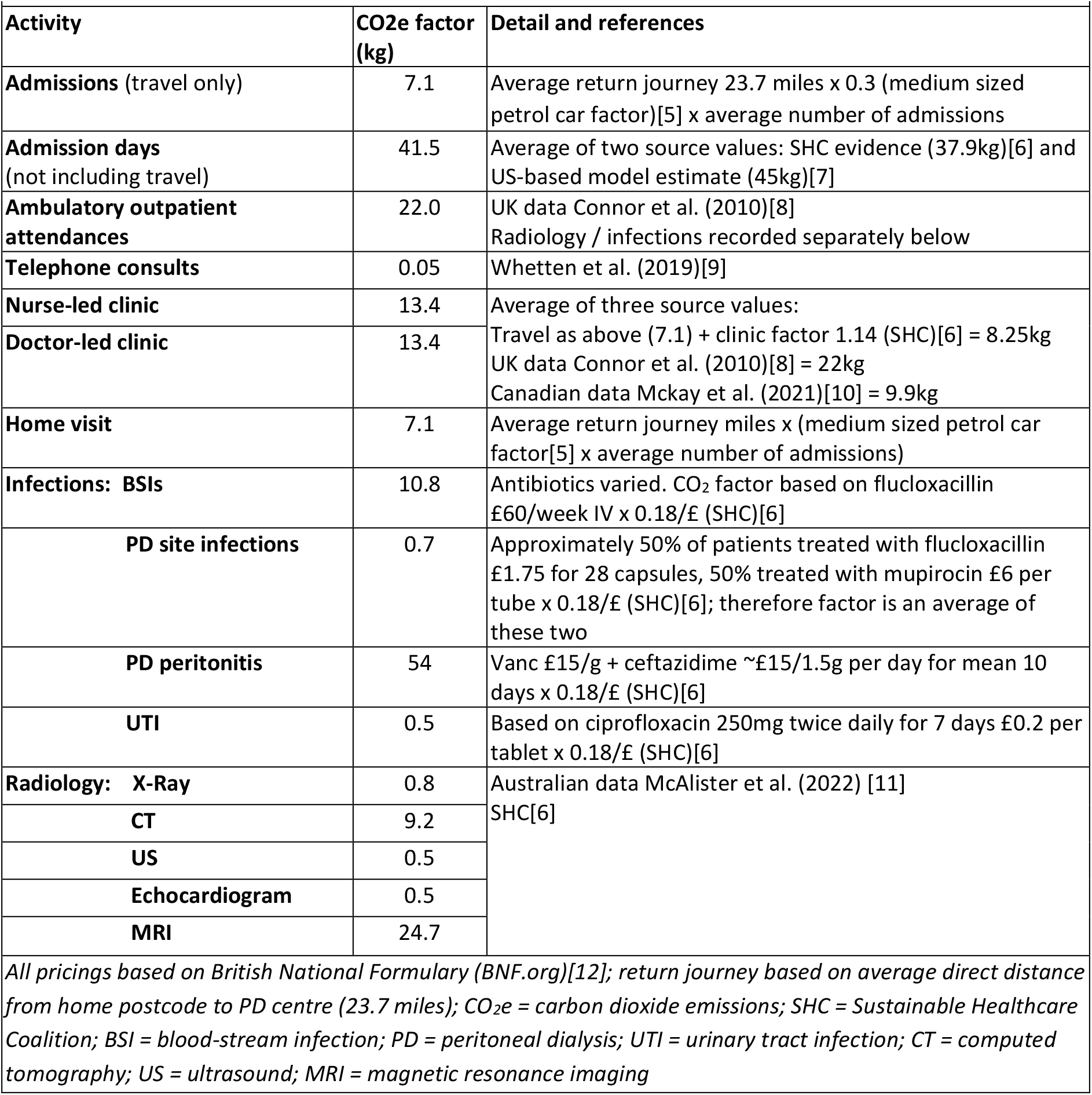
Factors utilised to calculate carbon mapping estimations for healthcare activity and their sources.

Previous studies have been undertaken in the same renal unit over the same period using consistent methodologies for patients on alternative modalities of KRT; the data were used for comparisons between modalities and references link to these data.

The study design, with use of routinely collected, anonymised patient data for the purposes of evaluating service performance for quality improvement, was classified as local service evaluation as opposed to research, therefore there was no requirement for review by an NHS research ethics committee.

## Results

PD was initiated in 122 patients between 2015 and 2019, totalling 30,976 days of PD. Table 2 reports the demographics of the study population. Seventy-two patients (59%) commenced automated peritoneal dialysis (APD) and the remainder continuous ambulatory peritoneal dialysis (CAPD).

**Table 2.**
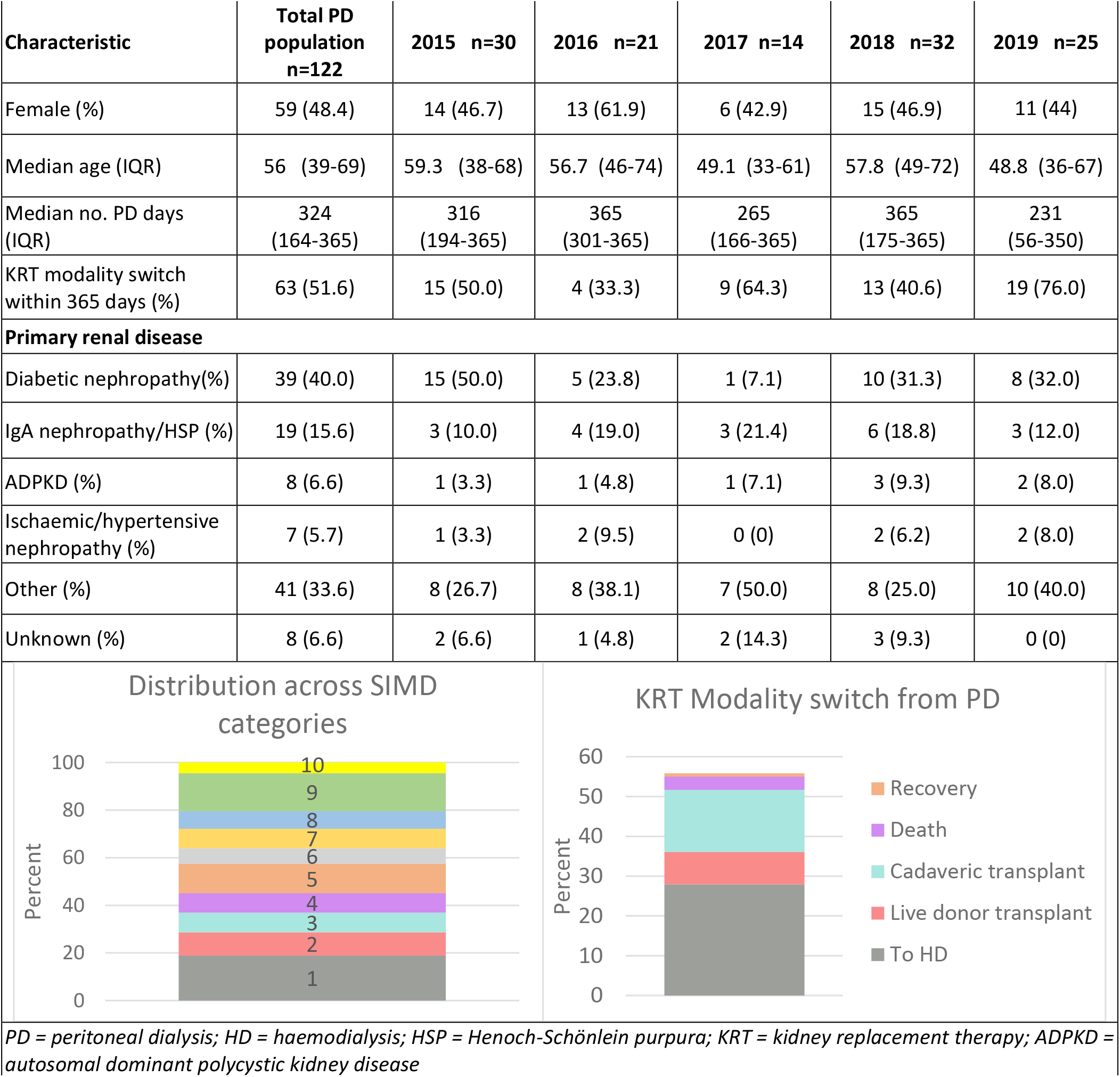
Demographics of incident PD population displayed by year commencing PD.

| Characteristic | Total PD population n=122 | 2015 n=30 | 2016 n=21 | 2017 n=14 | 2018 n=32 | 2019 n=25 |
| --- | --- | --- | --- | --- | --- | --- |
| Female (%) | 59 (48.4) | 14 (46.7) | 13 (61.9) | 6 (42.9) | 15 (46.9) | 11 (44) |
| Median age (IQR) | 56 (39-69) | 59.3 (38-68) | 56.7 (46-74) | 49.1 (33-61) | 57.8 (49-72) | 48.8 (36-67) |
| Median no. PD days (IQR) | 324 (164-365) | 316 (194-365) | 365 (301-365) | 265 (166-365) | 365 (175-365) | 231 (56-350) |
| KRT modality switch within 365 days (%) | 63 (51.6) | 15 (50.0) | 4 (33.3) | 9 (64.3) | 13 (40.6) | 19 (76.0) |
| **Primary renal disease** |  |  |  |  |  |  |
| Diabetic nephropathy(%) | 39 (40.0) | 15 (50.0) | 5 (23.8) | 1 (7.1) | 10 (31.3) | 8 (32.0) |
| IgA nephropathy/HSP (%) | 19 (15.6) | 3 (10.0) | 4 (19.0) | 3 (21.4) | 6 (18.8) | 3 (12.0) |
| ADPKD (%) | 8 (6.6) | 1 (3.3) | 1 (4.8) | 1 (7.1) | 3 (9.3) | 2 (8.0) |
| Ischaemic/hypertensive nephropathy (%) | 7 (5.7) | 1 (3.3) | 2 (9.5) | 0 (0) | 2 (6.2) | 2 (8.0) |
| Other (%) | 41 (33.6) | 8 (26.7) | 8 (38.1) | 7 (50.0) | 8 (25.0) | 10 (40.0) |
| Unknown (%) | 8 (6.6) | 2 (6.6) | 1 (4.8) | 2 (14.3) | 3 (9.3) | 0 (0) |

PD = peritoneal dialysis; HD = haemodialysis; HSP = Henoch-Schönlein purpura; KRT = kidney replacement therapy; ADPKD = autosomal dominant polycystic kidney disease

One patient had their care transferred to a neighbouring health board during the 365 day period studied. The median return distance travelled from a patient’s home to the renal unit was 11.7 (IQR 7.6-35.6) miles.

### Switching KRT Modality

Four patients died and 63 patients (51.6%) transitioned to another KRT within 365 days of commencing PD (10 live donor kidney transplantation, 19 cadaveric kidney transplantation, 34 haemodialysis) and one patient recovered native function. Figure 1 demonstrates a time series plot of patients continuing PD as a KRT modality throughout the 365-day period studied.

**Figure 1.**
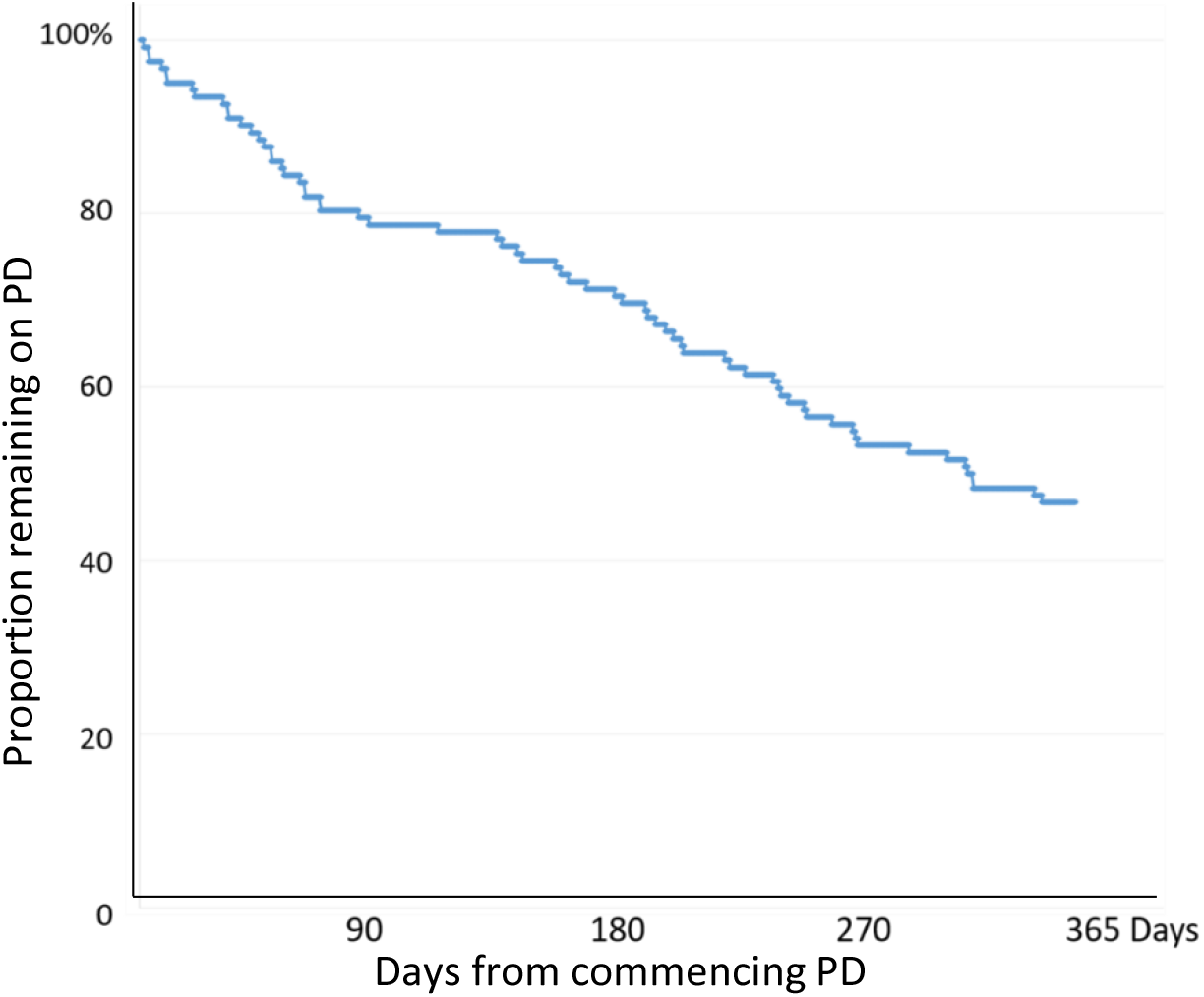
Kaplan-Meier estimator of PD over the incident year. See text for indications to discontinue PD.

There was a higher proportion of patients switching to HD within the first three months of commencing PD (17 patients, 50% of all those transitioning to HD within the first 365 days). Thereafter there was a stable proportion of patients switching to HD over the remaining three-month intervals of the year: 17.6%, 17.6% and 14.7% at 3-6 months, 6-9months and 9-12 months respectively.

Infection episodes were responsible for 35.3% of patients switching from PD to HD (8.8% recurrent peritonitis, 8.8% persistent peritonitis, 5.9% fungal peritonitis, 8.8% other peritonitis, 2.9% persistent abdominal wall cellulitis). Persistent catheter malfunction accounted for 8.8% of reasons for switching to HD and leak of peritoneal fluid outside the peritoneal cavity in 11.8% of cases.

Symptomatic solute clearance failure and failure of ultrafiltration accounted for 17.6% and 8.8% of cases necessitating switch in KRT. The remaining reasons for switch to HD from PD included patient choice/not aligning with lifestyle needs (8.8%), major abdominal surgery (5.9%) and symptomatic hypotension related to fluid exchange (2.9%).

### Dialysis access

The median duration from insertion of PD catheter to utilisation was 26 (IQR 17-50) days, with a range of 1 to 599 days. Eight patients (6.6%) required more than one insertion procedure throughout the 365 days. Ten patients (8.2%) required the insertion of a temporary non-tunnelled central venous catheter to enable haemodialysis due to complications preventing PD or PD inadequacy.

### Contact days with renal services

Over the first year, patients had a median 35 (IQR 23-45) days of face-to-face contact days with renal services. This included a total of 1147 admission days (37 in-patient days/1000 patient days) with 188 separate admission episodes (6 admissions/1000 patient days), and 1820 nurse-led (59 appointments/1000 patient days) and 855 doctor-led clinic appointments (28 appointments/1000 patient days), and 302 home visits (10 visits/1000 patient days).

The renal service contact days included a mean of 2.5 (SD 2.5) unscheduled ambulatory assessments per patient; 14.9 (SD 8.96) face-to-face clinics with a PD nurse and 7 (SD 3.9) with a nephrologist per patient; and 2.5 (SD 3.4) home visits from PD nurses per patient. Additionally patients had an average of 4.7 (SD 6.2) telephone consultations during the first year on PD.

There was a median of 1.0 (IQR 0-2) hospital admission and a median 5 (IQR 0-10.8) in-patient days. Figure 2 demonstrates admission day data across the 365 days visually and numerically. Admission days were variable throughout the year, highest in the first and the third quarter of the incident year, with additional clustering around KRT transitions (not included in Figure 2).

**Figure 2.**
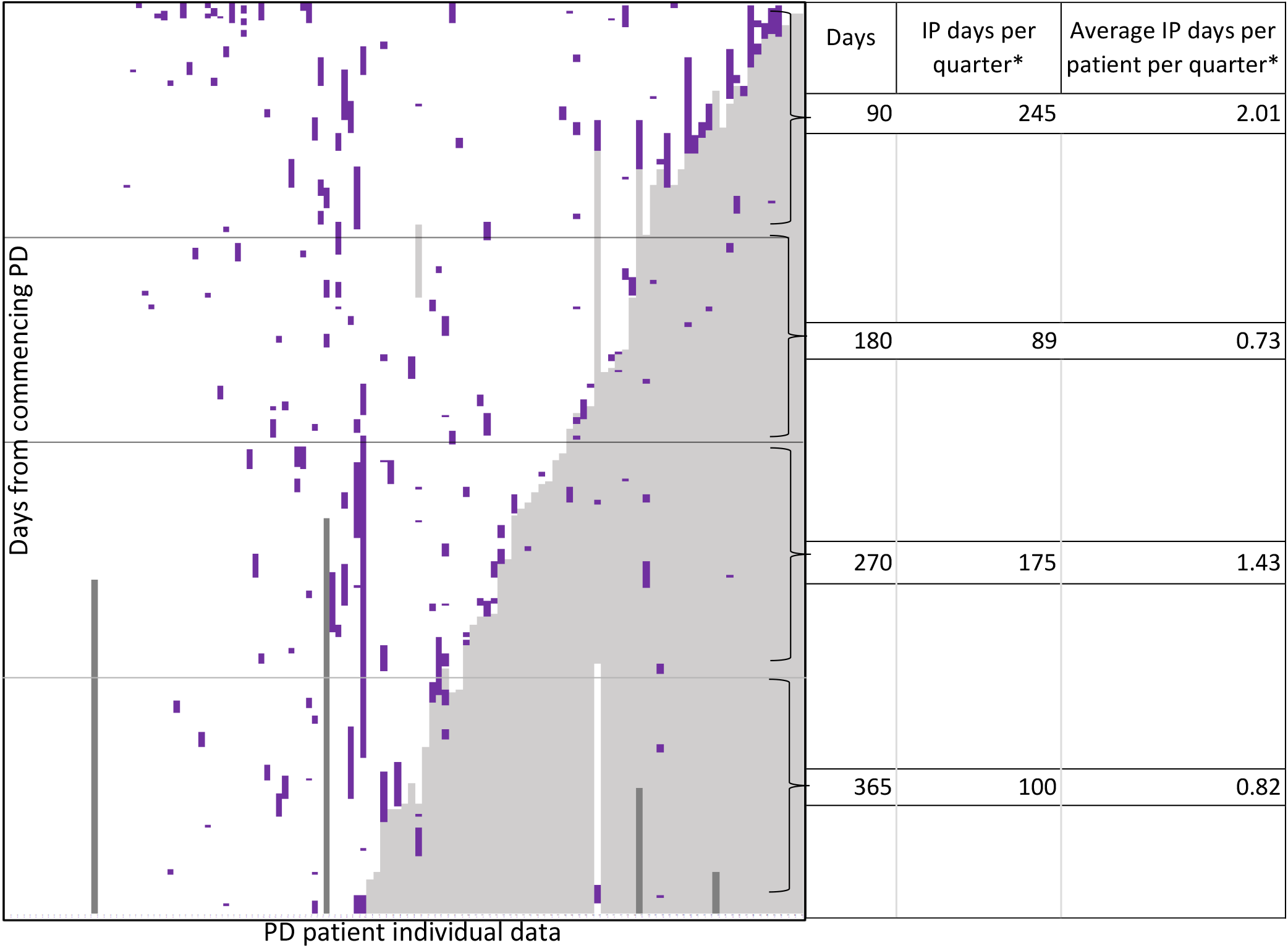
Inpatient admissions (purple boxes), kidney replacement therapy switch (light grey boxes) and death (dark grey boxes), with 90/180/270 day lines. Each column represents one patient. Day 0 at top of the rectangle and day 365 at the bottom. Adjacent table is quarterly admission days. *adjusted for censored days.

The most common reason for inpatient admission was infection episodes, followed by episodes related to vascular access complications or formation related to preparing for haemodialysis therapy. Table 3 illustrates a breakdown of primary reasons for inpatient admission.

**Table 3.** Primary reason for inpatient admission during first 365 days on PD.

|  |  |
| --- | --- |
| Reason for admission | Number of admissions |
| Infection | 61 |
| Vascular access (planned change to KRT) | 15 |
| Fluid overload | 14 |
| Cardiovascular | 13 |
| Surgery or injury | 17 |
| PD catheter malfunction | 10 |
| PD catheter removal | 10 |
| KRT transition related | 9 |
| PD catheter insertion/exchange | 9 |
| PD training | 9 |
| Gastrointestinal symptoms | 9 |
| Glycaemic disturbances | 4 |
| Uraemia or other metabolic encephalopathy | 4 |
| Electrolyte disturbance | 4 |
| *PD = peritoneal dialysis; KRT = kidney replacement therapy* |  |

Sixty-five patients (53.3%) had no admission days over the incident year. Eighteen of these admission-free patients completed the year on PD (27.7%). Forty-seven patients were admission-free up to the point of censoring for modality change, recovery of kidney function, or death which represents 72% of the 65 censored patients.

### Radiological activity and infection episodes

Over the first 365 days after starting peritoneal dialysis, on average there were 4.4 (SD 4.1) radiological investigations per patient, including 3.1/patient x-rays, 0.4/patient CTs, 0.4/patient ultrasounds and 0.1/patient MRIs. This equates to 17 investigations/1000 patient days.

There was a mean of 1.4 (SD 1.4) total infection events per patient over the year (5.6 per 1000 patient days). Figure 3 illustrates the pattern of infection episodes throughout the year. The average number of infection episodes per patient day was highest in the first quarter of the year (0.005) and reduced throughout each quarter with the lowest number of infection episodes occurring in the final quarter (0.002 average infections per patient day).

**Figure 3.**
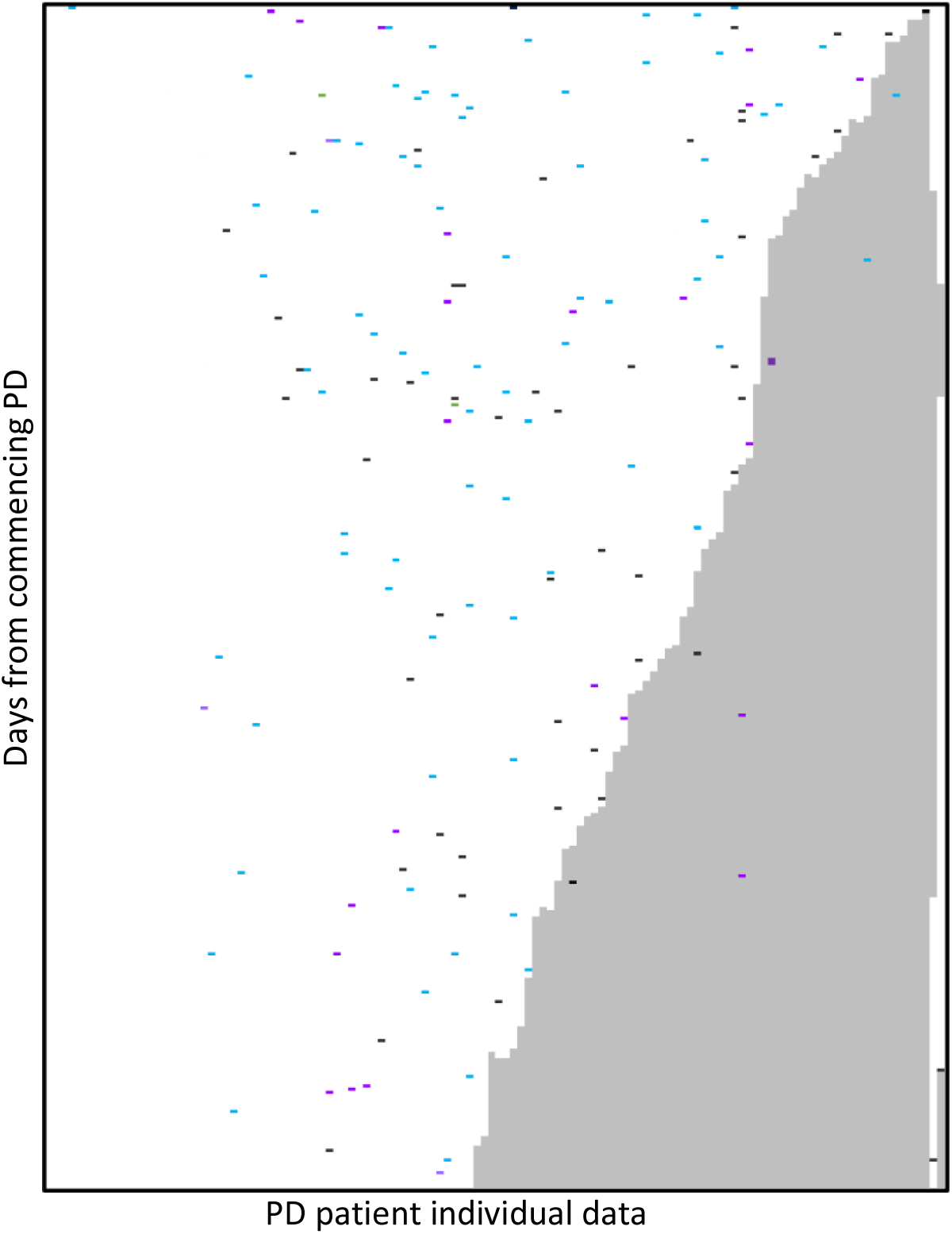
Infection episodes across the 365 day period. Day 0 at the top of the rectangle and day 365 at the bottom. Each column represents one patient, ordered by most ‘activity’ days left to least activity on right. Bacteraemia = green, UTI = purple, PD catheter exit site infection = blue, peritonitis = black, kidney replacement therapy transition/death censoring = light grey

There were 0.02/patient bloodstream infections, all of which were secondary to micro-organisms originating from skin contamination (two *Staphylococcus aureus*, one *Staphylococcus hominis* considered a contaminant result, one *Staphylococcus epidermidis* considered a contaminant result and one *Staphylococcus epidermidis* considered a pathological result). There were 0.2/patient urinary tract infections, 0.7/patient PD catheter site infections, and 0.5/patient PD peritonitis episodes.

There were 38 patients who experienced at least one episode of PD peritonitis during the year, and these patients had a PD catheter in situ for a median of 30 days prior to use. There were instances of infection events in the time period between PD catheter insertion and utilisation, with an average of 0.2 (SD 1.2) PD catheter site infection episodes per patient and 0.05 (SD 0.3) PD peritonitis episodes.

#### Carbon mapping

The estimated median carbon footprint from patient in- and out-patient contact with renal services (but excluding the peritoneal dialysis itself) was 581 kg CO2e/patient over the incident year of PD. This included a median 207 kg CO2e/patient for inpatient days, 55 kg CO2e/patient for unscheduled ambulatory assessment, 0.2 kg CO2e/patient for telephone consultations, 294 kg CO2e/patient for outpatient clinic activity and 18 kg CO2e/patient for home visits. The total for the year was inclusive of 9 kg CO2e/patient for radiological investigation and 26 kg for treatment of infection episodes.

## Discussion

This study illustrates the associated healthcare burden on patients commencing peritoneal dialysis for end-stage kidney disease and highlights the carbon footprint associated with this. This information has dual utility; firstly for service providers, in anticipating care requirements and planning carbon reduction strategies. Secondly, such data can inform patient decision making when receiving education on kidney replacement therapies. Patients should be aware of the frequency of attendance and admission days in the first year, despite PD being described as a ‘home-based’ therapy.

The median 5 hospitalised bed days in the first year of PD in our study compares to 18.1 days for a contemporary incident HD cohort, with 1.0 versus 2.0 admissions respectively in the first 365 days of initiating therapy[3]. During the first two years of a contemporary transplant cohort, patients had a median number of 47 days clinical contact which included 15 in-patient days[4]. Considering admission days per 1000 KRT days for each modality, PD is associated with lower rates than HD (admission days 37/1000 vs 51.6/1000 KRT days for PD and HD respectively, higher hospital admission days than transplant allocation category R1-3 (27.4/1000) and lower than R4 transplant recipients (47.1/1000)[3][4].

There is variability across published studies regarding the relative rate of hospitalisation between KRT modalities. Recent European studies of incident dialysis patients have noted a higher rate of hospital admission and longer length of stay in patients undergoing peritoneal dialysis compared with those on haemodialysis[13][14]. It is notable the median age of patients in these cohorts was between 6-9 years older than our cohort. Similar to our study the most common reason for admission in patients undergoing PD in these studies is infection, specifically peritonitis[13][14]. It may be that we attain lower hospital admission days lower due efficacy of same day assessment and supported ambulatory or home delivery of antibiotics to allow the patient to avoid admission.

KRT transition was common in this cohort, 51.6%, with equal rates of transplantation and conversion to HD. Comparing this cohort to a haemodialysis cohort in the same centre where 81% of patients remained on haemodialysis at the end of 1 year, versus the 45% remaining on peritoneal dialysis in this study.[3] The proportion of patients remaining on PD and HD after 1 year according to the most recently published UK renal registry data is comparable at 58.6% and 73.2% respectively[1]. In a Dutch study of incident haemodialysis and peritoneal dialysis patients followed up on average for 22 months, there was a higher rate of conversion of patients from PD to HD versus converting from HD to PD, which is concurrent with UK-wide trends[1][14]. Additionally patients undergoing PD had a shorter dialysis vintage than those commencing HD[14]. The international Peritoneal Dialysis Outcomes and Practice Patterns Study (PDOPPS) of more than 2500 patients did not demonstrate an association with functional status impairment and transfer from PD to HD. The UK was one of the countries studied where patients utilising PD were demonstrated to have a lower functional score compared with other regions[15].

Infections were the most common reason for switching modality to haemodialysis, and given the implications of this and the impact on inpatient admission rates, requires a quality improvement focus in line with the Kidney Disease Outcomes Quality Initiative and the International Society for Peritoneal Dialysis guidance[16][17]. Sixty-eight percent of our cohort experienced at least one infection episode necessitating anti-microbials, with implications for patient experience and anti-microbial stewardship. This is compared to a contemporaneous transplant cohort where 48.5% experienced at least one clinically significant infection and 8.8% experienced 10+ infections over a two-year period[4]. Bloodstream infection incidence for patients on PD was much lower compared with that for patients on haemodialysis (0.54 vs PD 0.055/1000 days)[3]. The length of time PD catheters are in situ but not used was heterogeneous and merits further examination given the frequency of infection episodes and rate of KRT transition.

### Environmental impact

Combining analyses of patient-related outcomes and patient experience with consideration of resource utilisation and environmental footprint remains novel, though such focus will be increasingly common as we move into the era of ‘Net Zero’ carbon emissions targets. Whilst carbon equivalents do not fully capture the environmental impact of healthcare activity, as an available and widely understood metric they serve a role in optimising high value service delivery. Our data only relate to healthcare activity that arises parallel to the delivery of PD, not the green-house gas emissions intrinsic to the PD itself, but do demonstrate carbon hotspots relating to patient travel, hospital admissions, and episodes of peritonitis.

Existing data based on a Chinese study of 68 patients undergoing PD, reports the annual patient carbon footprint at ∼1.4t CO_2_e, assuming a daily dialysate dose of eight litres[18]. Variability arises from the number of exchanges, differences between APD and CAPD in the carbon embedded in the manufacture of the APD machine and its energy use; and drain method. Home PD requires that PD fluids are packaged in plastic and transported long distances, though patient transport is then minimal[18]. Packaging is an issue in all areas of kidney care, and was not analysed specifically in our study, but has been estimated as the dominant source of greenhouse gases in PD therapy (80%), suggesting that reductions are likely achievable with through collaboration with manufacturers and focus on waste management[18]. Globally, annual PVC plastic use in PD delivery is estimated at 30 000 metric tons and not currently recycled[19].

A full life cycle analysis of PD to include dialysis itself and pharmaceuticals is challenging, though currently being undertaken through a collaboration between UK Kidney Association and Sustainable Healthcare Coalition. The data provided by such research will benchmark these tariffs to permit more commonplace consideration of environmental sustainability as an aspect of healthcare. In the meantime, the environmental impact of PD may be reduced through wider use of virtual appointments, facilitated by electronic transfer of data from APD machines. As remote living is often an influencing factor in the decision to choose PD, road-miles saved can be significant. Working with procurement to reduce packaging and minimise plastic, and optimising delivery schedules is another approach to lessen environmental impact. Waste can also be reduced through effluent drain systems that keep consumables to a minimum and recycling.

Having identified hospital admissions and peritonitis as additional carbon-hotspots in our study, use of day-wards or ambulatory care systems can benefit both patient experience and service provider, with intra-peritoneal antibiotics likely to have a lower carbon footprint than intravenous. Finally, physical activity is an under-used, planet-friendly component of health and quality of life. Exercise should be promoted in individuals affected by CKD to delay the need for KRT, and following PD start should be continued to reduce the risk of co-morbidities[20].

Our study has limitations, with its small single-centre population and retrospective observational nature over a short period of time. The data was prospectively recorded although not designed for use in this study. Furthermore patient-specific factors that may influence choice of KRT were not included such as frailty scoring or employment, nor were patient-reported outcome data examined. Furthermore, the contribution of physician bias to KRT modality choice was not explored; which may be relevant as our incident rates of PD are lower than the UK average at 12.2% possibly reflecting higher rates of pre-emptive transplant, or a more conservative approach to referrals for PD training[21]. Patient demographics are however similar to the rest of the UK, supporting applicability to other centres.

This study illustrates the burden of kidney-associated healthcare on patients commencing peritoneal dialysis in the first year of using the modality, particularly with regard to contact with renal services and incidence of infection. Furthermore it highlights the carbon footprint associated with healthcare activity. Estimates of carbon footprint relating to routine and unscheduled care indicate that carbon hotspots include patient travel and hospital admissions, and episodes of peritonitis; a full life cycle analysis is merited.

## Data Availability

The data generated and analysed during the current study are not publicly available, because informed consent to share data for secondary use beyond this analysis was not obtained from participants.

## Acknowledgements

This research did not receive any specific grant from funding agencies in the public, commercial, or not-for-profit sectors.

## Authors’ Contributions

Kirsty Crowe, Peter Thomson and Eleanor Murray contributed to the conception and design of the study, and James Traynor and Joan MacLeod contributed to the analysis and interpretation of the data. Kirsty Crowe and Eleanor Murray created the original drafts of the manuscript and Peter Thomson, James Traynor and Joan MacLeod contributed to manuscript revision. All authors approved the submitted version and agree to be accountable for all aspects of the work.

